# Long-Term Recall of Amyloid PET Results

**DOI:** 10.1101/2025.04.10.25325602

**Authors:** Maria Scanlan, Hunter Heigert, Ryan A. Townley, Abdulaziz Almahgraby, Mickeal N. Key, Jill K. Morris, Jeffrey M. Burns, Eric D. Vidoni

**Affiliations:** University of Kansas Alzheimer’s Disease Research Center, University of Kansas Medical Center, Department of Neurology, Fairway, KS

**Author notes:** Corresponding Author: Eric D. Vidoni, PhD, University of Kansas Alzheimer’s Disease Research Center.

## Abstract

We assessed long-term recall and perceived impact of amyloid PET results among 92 cognitively normal adults who received structured result disclosure 6–9 years earlier. Two-thirds correctly recalled their result; only three misremembered the result as the opposite of what was communicated. Participants reported limited behavior change, though those with elevated scans more often endorsed lifestyle modifications. Perceived AD risk aligned with scan status but did not appear to cause lasting psychological distress. Most participants, regardless of result, viewed the experience positively and would undergo testing again. These findings support the long-term acceptability of disclosing amyloid PET results.

## Introduction

Alzheimer’s disease (AD) is among the most feared and stigmatized conditions globally ^1-5^. Recent scientific and clinical advances have opened a window of opportunity to treat and potentially prevent cognitive decline due to AD. However, a cure remains elusive. Notably, a growing number of biomarker tests for AD are clinically available or soon will be, creating a situation where clinicians can communicate disease risk well before symptom onset. This has raised concerns about the psychological ramifications of disclosing risk information to asymptomatic individuals.

The REVEAL study was the first to systematically assess the psychological impact of learning APOE ε4 carrier status. Results of that seminal study demonstrated little negative psychological impact ^6-8^. More recently, we and others have investigated newer AD risk markers such as amyloid positron emission tomography (PET) scans ^9-11^. These studies have shown that disclosing amyloid PET results is safe and well-tolerated. However, there is limited research on the long-term recall or outcome of learning AD biomarker results. Does learning one’s amyloid PET status leave a lasting imprint, as might be expected given the pervasive stigma surrounding dementia? In the present study, we evaluated long-term recall of cerebral amyloid status among individuals informed of their result several years earlier. We hypothesized that elevated amyloid status would be more memorable and would lead to greater health behavior adoption.

## Methods

We selected 100 individuals who received an 18F-AV-45 PET scan as part of screening for the APEx study (NCT02000583),^12^ which investigated the effects of exercise on cerebral amyloid load in individuals without cognitive impairment. During the APEx study screening, we informed all participants of their amyloid PET results through a structured disclosure process, that included telling participants they had “elevated” or “not elevated” cerebral amyloid.^10^ The process and information were shown to be safe with minimal sustained anxiety or depression about the result.

For the current study, we used a pseudo-random selection method to balance participants who enrolled in APEx with those who screen-failed, aiming to enrich the sample with individuals who had an “elevated” scan. Participants were contacted 6 to 9 years post-scan.

This study was approved by the University of Kansas Medical Center Institutional Review Board, which allowed the use of verbal consent obtained over the phone. Our questionnaire assessed participants’ recall of their PET result, perceived AD risk, emotional response to the disclosure, and any behavior changes following the result.

Participants were asked to categorize their recalled PET result as “Elevated,” “Not Elevated,” or “Don’t Remember.” They also rated their current perceived risk of developing AD as “No Risk,” “Low Risk,” “Moderate Risk,” or “High Risk.” Emotional response to the scan result was rated as “Positive,” “Neutral,” or “Negative.”

Participants were also asked whether they had taken specific actions following the disclosure: discussed results with family, consulted a doctor, made lifestyle changes, began vitamins or supplements, or reviewed estate plans. They were also asked whether they would choose to receive the scan again. Responses were dichotomized as “Endorsed” or “Not Endorsed.”

We calculated frequencies and proportions for categorical variables. Between-group comparisons (“Elevated” vs. “Not Elevated”) were made using Pearson’s chi-squared test or Fisher’s exact test for categorical variables, and Welch’s t-test for continuous variables. Significance was defined as p < 0.05. Agreement between communicated and recalled results was assessed using percent agreement.

## Results

Of the 100 individuals selected, 94 were reached, and 92 consented to participate. The mean age of participants was 78.5 years (SD = 4.4); two-thirds (n = 62) were female.

In general, participants remembered the results of their 18F-AV-45 PET scan, with 66% agreement between the result communicated to them and the result they recalled 6-9 years later. Of note, only 3 participants recalled the opposite result of what they had been told their scan results. And almost one third (n=28 of 92) could not remember their results, with similar frequencies of forgetting between those who were “Elevated” and those who were “Not Elevated”. The groups did not differ in their accuracy of their recollection (p = 0.8). Through our longitudinal cohort study, we have a research evaluation^13^ on 34 individuals. At their last known visit, six individuals had experienced cognitive change. Their recall accuracy did not significantly differ from those who remained cognitively stable (p = 0.6). Additionally, recall accuracy among individuals with unknown follow-up status was not significantly different from those who remained cognitively normal (p = 0.8).

Learning the PET scan results did little to change behaviors, per participant report. We identified no differences in participant reports of discussing results with family or physician, insurance, estate, or asset changes or reallocation, or taking vitamins or supplements (p >= 0.2). The only area where people did report a change was in lifestyle behaviors (p < 0.001). Participants who were “Elevated” reporting adopting one or more health behavior changes such as better diet and exercise. It is possible that this reflects their participation in the APEx exercise study itself, and we have no data regarding if participants sustained these behavior changes.

Perceptions of AD risk differed significantly by group (p < 0.001). Participants with an “Elevated” scan were more likely to report moderate to high perceived risk (88%). However, participants in the “Not Elevated” group appeared to recognize and remember learning from the counseling at the time of the scan that scans were not deterministic. More than 40% rated themselves as having moderate to high risk for developing AD. Emotional reactions differed as well: “Not Elevated” participants tended to report positive feelings (81%), while those in the “Elevated” group more often reported neutral (48%) or negative (22%) reactions. Nevertheless, 88% of all participants said they would choose to undergo the scan again.

## Discussion

We were among the first to test the safety of amyloid PET result disclosure in cognitively normal research participants, finding it to be safe and well-tolerated.^10^ In this follow-up, we observed that recall of PET scan results remains relatively strong years later, but not perfect. The result disclosure had limited long-term impact on behavior or psychological distress.

These findings are consistent with prior research. Lingler et al.^14^ found that individuals with mild cognitive impairment (MCI) emphasized the benefits over harms of knowing their amyloid PET results. Other work with individuals who have Mild Cognitive Impairment has found good short-term recall of PET results.^11^, and like the REVEAL study,^7^ we found a reported increase in healthy behavior—though we could not verify execution. Another limitation is the lack of information on verified cognitive impairment in a majority of the sample.

Given the fear and stigma that still surrounds AD, it is reasonable to assume that individuals would place significant emotional weight on test results and that they would be highly memorable. Instead, we found that even in the long term, haring the results of an AD risk test had relatively little impact on behavior, psychological distress, and were not indelible. Despite widespread public concern surrounding AD, our findings suggest that disclosing AD biomarker information does not lead to lasting psychological harm or major life changes. Many individuals find the information worthwhile, even years later. Though not all remember their findings.

## Data Availability

Reasonable data requests will be accommodated by request to the corresponding author.

## Availability of Data

Reasonable data requests will be accommodated by request to the corresponding author.

## Funding

This work was supported by P30 AG072973.

**Table.**

|  | Overall<br>N = 92<sup>1</sup> | Not Elevated<br>N = 65<sup>1</sup> | Elevated<br>N = 27<sup>1</sup> | Sig.<sup>2</sup> |
| --- | --- | --- | --- | --- |
| **Recollection of Status** |  |  |  |  |
| Elevated | -- | 2 / 65 (3.1%) | 19 / 27 (70%) | -- |
| Not Elevated | -- | 42 / 65 (65%) | 1 / 27 (3.7%) | -- |
| Does Not Remember | -- | 21 / 65 (32%) | 7 / 27 (26%) | -- |
| **Discussed With Family** | 51 / 92 (55%) | 36 / 65 (55%) | 15 / 27 (56%) | >0.9 |
| **Talked With Doctor** | 22 / 92 (24%) | 15 / 65 (23%) | 7 / 27 (26%) | 0.8 |
| **Changed Insurance** | 1 / 92 (1.1%) | 1 / 65 (1.5%) | 0 / 27 (0%) | >0.9 |
| **Changed Assets** | 0 / 92 (0%) | 0 / 65 (0%) | 0 / 27 (0%) | >0.9 |
| **Changed Lifestyle** | 28 / 92 (30%) | 12 / 65 (18%) | 16 / 27 (59%) | <0.001 |
| **Began Vitamins or Supplements** | 10 / 92 (11%) | 5 / 65 (7.7%) | 5 / 27 (19%) | 0.2 |
| **Reviewed Estate** | 4 / 92 (4.3%) | 2 / 65 (3.1%) | 2 / 27 (7.4%) | 0.6 |
| **Changed Nothing** | 19 / 92 (21%) | 15 / 65 (23%) | 4 / 27 (15%) | 0.4 |
| **Perceived Risk of AD** |  |  |  | <0.001 |
| No risk | 8 / 87 (9.2%) | 8 / 62 (13%) | 0 / 25 (0%) |  |
| Low risk | 31 / 87 (36%) | 28 / 62 (45%) | 3 / 25 (12%) |  |
| Moderate risk | 40 / 87 (46%) | 24 / 62 (39%) | 16 / 25 (64%) |  |
| High risk | 8 / 87 (9.2%) | 2 / 62 (3.2%) | 6 / 25 (24%) |  |
| **Emotional Impact of PET** |  |  |  | <0.001 |
| Positive | 50 / 76 (66%) | 43 / 53 (81%) | 7 / 23 (30%) |  |
| Neutral | 21 / 76 (28%) | 10 / 53 (19%) | 11 / 23 (48%) |  |
| Negative | 5 / 76 (6.6%) | 0 / 53 (0%) | 5 / 23 (22%) |  |
| **Scan Again** | 73 / 83 (88%) | 54 / 60 (90%) | 19 / 23 (83%) | 0.5 |

<sup>1</sup>Mean ± SD; n / N (%)<sup>2</sup>Welch Two Sample t-test; Pearson's Chi-squared test; Fisher's exact test

